# Natural Language Processing for Adjudication of Heart Failure Hospitalizations in a Multi-Center Clinical Trial

**DOI:** 10.1101/2023.08.17.23294234

**Authors:** Jonathan W. Cunningham, Pulkit Singh, Christopher Reeder, Brian Claggett, Pablo M. Marti-Castellote, Emily S. Lau, Shaan Khurshid, Puneet Batra, Steven A. Lubitz, Mahnaz Maddah, Anthony Philippakis, Akshay S. Desai, Patrick T. Ellinor, Orly Vardeny, Scott D. Solomon, Jennifer E. Ho

**Affiliations:** Division of Cardiovascular Medicine, Brigham and Women’s Hospital, Boston, Massachusetts; Cardiovascular Disease Initiative, Broad Institute of Harvard University and the Massachusetts Institute of Technology, Cambridge, Massachusetts; Data Sciences Platform, Broad Institute of Harvard and the Massachusetts Institute of Technology, Cambridge, Massachusetts; Division of Cardiology, Massachusetts General Hospital, Boston, Massachusetts; Demoulas Center for Cardiac Arrhythmias, Massachusetts General Hospital, Boston, Massachusetts; Minneapolis VA Hospital, University of Minnesota, Minneapolis, Minnesota; CardioVascular Institute and Division of Cardiology, Department of Medicine, Beth Israel Deaconess Medical Center, Boston, Massachusetts

## Abstract

**Background:** The gold standard for outcome adjudication in clinical trials is chart review by a physician clinical events committee (CEC), which requires substantial time and expertise. Automated adjudication by natural language processing (NLP) may offer a more resource-efficient alternative. We previously showed that the Community Care Cohort Project (C3PO) NLP model adjudicates heart failure (HF) hospitalizations accurately within one healthcare system.

**Methods:** This study externally validated the C3PO NLP model against CEC adjudication in the INVESTED trial. INVESTED compared influenza vaccination formulations in 5260 patients with cardiovascular disease at 157 North American sites. A central CEC adjudicated the cause of hospitalizations from medical records. We applied the C3PO NLP model to medical records from 4060 INVESTED hospitalizations and evaluated agreement between the NLP and final consensus CEC HF adjudications. We then fine-tuned the C3PO NLP model (C3PO+INVESTED) and trained a *de novo* model using half the INVESTED hospitalizations, and evaluated these models in the other half. NLP performance was benchmarked to CEC reviewer inter-rater reproducibility.

**Results:** 1074 hospitalizations (26%) were adjudicated as HF by the CEC. There was high agreement between the C3PO NLP and CEC HF adjudications (agreement 87%, kappa statistic 0.69). C3PO NLP model sensitivity was 94% and specificity was 84%. The fine-tuned C3PO and *de novo* NLP models demonstrated agreement of 93% and kappa of 0.82 and 0.83, respectively. CEC reviewer inter-rater reproducibility was 94% (kappa 0.85).

**Conclusion:** Our NLP model developed within a single healthcare system accurately identified HF events relative to the gold-standard CEC in an external multi-center clinical trial. Fine-tuning the model improved agreement and approximated human reproducibility. NLP may improve the efficiency of future multi-center clinical trials by accurately identifying clinical events at scale.

## Introduction

Heart failure is the leading cause of hospitalization in the United States.^1^ Accurate identification and classification of heart failure events is challenging, due to clinical and phenotypic heterogeneity, as well as the absence of definitive and scalable diagnostic testing. Yet, the validity of heart failure randomized trials, epidemiology, and outcomes research depends on accurate identification of heart failure hospitalization events. The gold-standard for defining HF outcomes is adjudication by a randomized trial clinical events committee (CEC), a time-intensive process in which expert physicians manually review medical records and determine whether the case meets established criteria.^2^ For studies too large for practical CEC adjudication, *International Classification of Disease* (ICD) codes are commonly used to define heart failure hospitalizations. Of note, ICD codes for heart failure are known to be imprecise: approximately 40% of hospitalizations with primary position ICD codes for heart failure do not meet established criteria on medical record review.^3–6^ Automated adjudication of heart failure hospitalizations at scale may provide a more resource-efficient alternative or complementary approach to CEC adjudication that could enable larger and less expensive clinical trials and improve the quality of epidemiology and health policy research.

Natural language processing (NLP) is a potential strategy for accurate and automated clinical outcome adjudication. Innovations in pre-trained transformer-based NLP architectures have enabled the development of accurate models for specific classification tasks requiring only modest numbers of expert-labeled training examples. In prior work, we developed and internally validated a transformer-based NLP model which accurately adjudicates heart failure hospitalizations from discharge summary text in a single-center electronic health record cohort, the Community Care Cohort Project (C3PO).^6^ Since future applications of large-scale NLP adjudication at scale would include many centers, it is essential to evaluate whether the accuracy of this model generalizes beyond a single center, before considering implementation at scale. Indeed, machine learning models can be overfit to a particular care setting and thus generalize poorly.^7–9^ In this study, we externally validated the C3PO NLP model outside the healthcare system in which it was developed and compared to a gold-standard CEC in a multi-center NIH-sponsored clinical trial, INVESTED (Influenza Vaccine to Effectively Stop Cardio Thoracic Events and Decompensated Heart Failure).^10, 11^

## Methods

### Patients and Clinical Adjudication

The design and primary results of the INVESTED trial have been published.^10, 11^ Briefly, INVESTED enrolled 5,260 patients with recent acute myocardial infarction or heart failure hospitalization and at least one additional risk factor at 157 sites in the United States and Canada from September 2016 through January 2019. Participants were randomized to high-dose trivalent influenza vaccine or standard-dose quadrivalent influenza vaccine. High dose compared to standard dose influenza vaccine did not significantly reduce the incidence of the primary outcome (all-cause death or hospitalization for cardiac or pulmonary reason) or any key secondary outcome. INVESTED was sponsored by the National Heart, Lung, and Blood Institute. Institutional review boards at each site approved the study protocol. All patients provided written informed consent.

INVESTED sites submitted medical records (admission note or discharge summary at minimum) for all deaths and hospitalizations considered by the site investigator as potentially for a cardiopulmonary reason for adjudication by the central CEC at the Brigham and Women’s Hospital Clinical Trials Outcome Center (Boston, USA). The CEC included 21 physicians. The cause of each hospitalization was adjudicated by the CEC according to pre-specified criteria which were based on the 2017 Cardiovascular and Stroke Endpoint Definitions for Clinical Trials, written by the Standardized Data Collection for Cardiovascular Trial Initiative (SCTI) and the United States Food and Drug Administration (FDA).^2^ Each hospitalization dossier was adjudicated independently by two CEC physician reviewers. When the two reviewers agreed, the adjudication was considered final. When the two reviewers did not agree, the case was discussed in a meeting of the full CEC, with the CEC chair serving as the tiebreaker.

### Preparation of INVESTED Dossiers for NLP Adjudication

Adjudication dossiers were converted from Portable Document Format to TIFF images files using Ghostscript Seamless^12^ and from TIFF to text using Tesseract optical character recognition.^13^ The quality of optical character recognition was high. In order to assess NLP performance in adjudication medical records alone, a cover sheet describing the site investigator’s adjudication of the hospitalization’s cause and a brief investigator summary of the event was removed prior to NLP inference. Hospitalizations in INVESTED were defined as unplanned admissions to an acute care facility; elective procedures, emergency department visits, and outpatient visits submitted to the CEC (n=364) were not adjudicated and therefore excluded from this analysis. Dossiers with no available medical records beyond the cover sheet (n=270), or with medical records in French (n=142) were excluded. Due to the primary endpoint of hospitalization for primary cardiovascular or pulmonary reason, the INVESTED CEC event definitions included a “cardiopulmonary hospitalization (non-specific)” category for hospitalizations” that could be secondary to either congestive heart failure or pulmonary etiology but for which the primary etiology cannot be determined.” The 300 hospitalizations (7%) that received a final CEC adjudication of “cardiopulmonary non-specific” were excluded from the primary NLP validation because the CEC did not provide a gold-standard adjudication of heart failure, but were included in a sensitivity analysis.

### External Validation of the C3PO Heart Failure NLP Model

The development and validation of the C3PO NLP model for heart failure hospitalization has been previously described.^6^ Briefly, cardiologists adjudicated 1934 discharge summaries from hospitalizations with ICD codes for heart failure in C3PO, an electronic health record cohort of patients receiving longitudinal primary care at Mass General Brigham.^14^ Heart failure hospitalizations were defined using commonly accepted criteria employed by clinical trial CECs.^2^ Multiple transformer-based NLP models based on different pre-training architectures were developed in a training set (n=1268), and their performance was compared in a test set (n=214). The best model was based on Clinical Longformer pre-training.^15^ Since the NLP model produces a continuous score reflecting likelihood of heart failure, a threshold for binary adjudication of heart failure hospitalization was defined in the test set. In a held-out internal validation set, adding the C3PO NLP heart failure model to ICD codes improved adjudication accuracy compared to the gold standard of clinician review.

The study design of this validation study is outlined in **Figure 1**. The text from each INVESTED medical record dossier was submitted to the C3PO NLP model. The previously published threshold for converting continuous NLP heart failure score to a binary heart failure adjudication in the C3PO model was used without recalibration. Agreement between the C3PO NLP heart failure adjudication and the final CEC adjudication (heart failure vs non-heart failure) was assessed by Cohen’s kappa statistic, sensitivity, specificity, and positive and negative predictive value, using the CEC adjudication as the gold standard for test characteristics. C3PO NLP model kappa was assessed in key subgroups. Inter-rater reproducibility between the two independent human reviewers on the INVESTED CEC was assessed on the same set of hospitalizations. Final CEC adjudications were tabulated across each decile of continuous C3PO NLP score.

**Figure 1:**
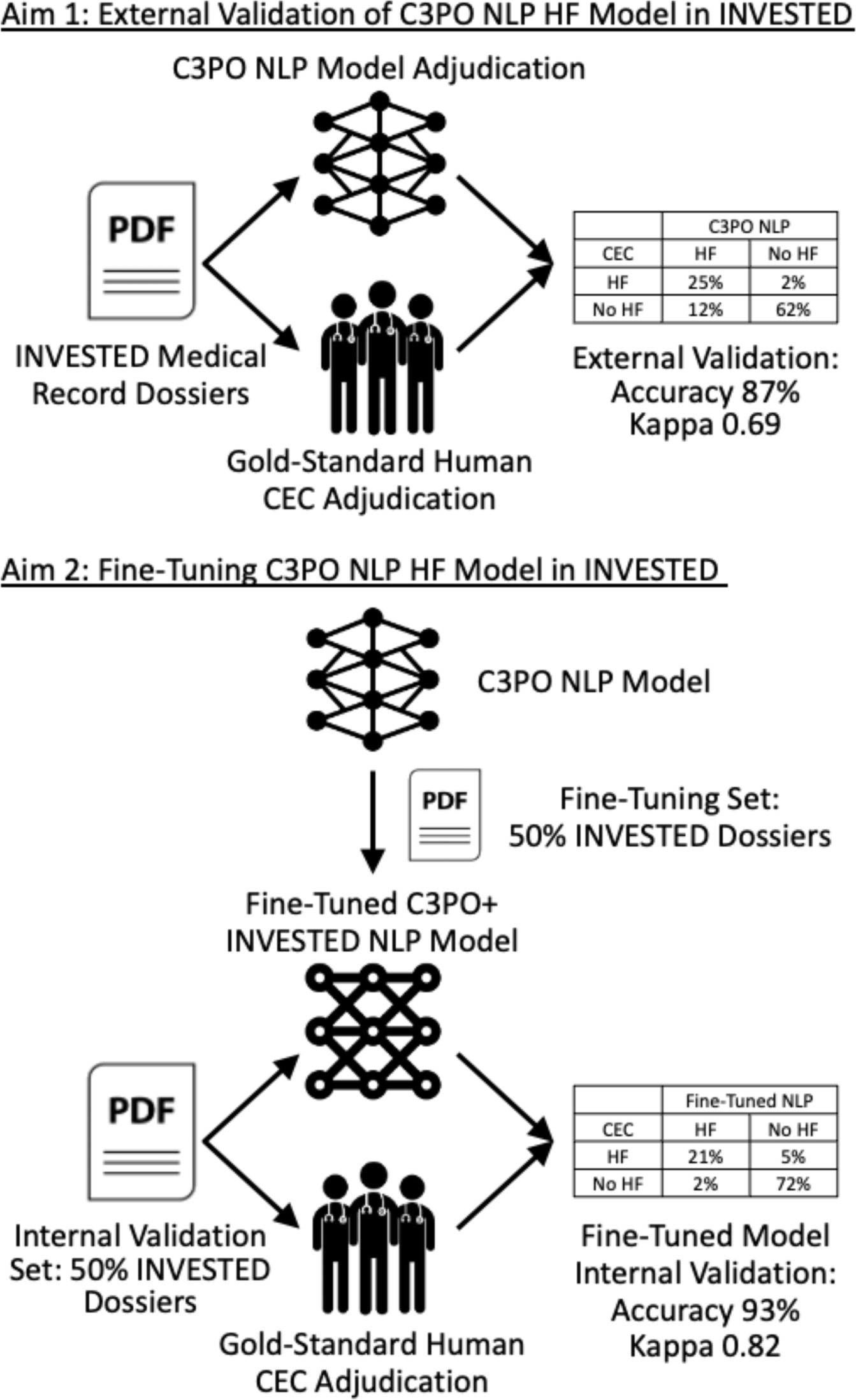
Study Design. CEC, clinical event committee; C3PO, Community Care Cohort Project; HF, heart failure; NLP, natural language processing

### Fine-Tuning the C3PO NLP Model and Training a *De Novo* NLP Model in INVESTED

We then developed and assessed new NLP models created by a) fine-tuning the C3PO NLP model, and b) training a new model, using a subset of INVESTED records. We separated INVESTED hospitalizations randomly and approximately evenly into a development set (1,954 hospitalizations) and internal validation set (2,106 hospitalizations), ensuring that there were no overlapping individuals between sets. The fine-tuned C3PO+INVESTED model was created by initializing Clinical Longformer with the weights from the C3PO NLP model and further training it on labeled dossiers from the INVESTED development set. Up to 400 hospitalizations from the development set were used for hyperparameter optimization. The *de novo* INVESTED NLP model was created by training Clinical Longformer in the development set without using the C3PO model weights. These models were evaluated in the held-out internal validation set, against the gold-standard INVESTED CEC adjudication, as in the primary validation analysis. To investigate whether a smaller development set would have been sufficient, we trained versions of the C3PO+INVESTED and INVESTED NLP models using subsets of the development set, and evaluated them on the identical validation set.

## Results

### Baseline Characteristics

A total of 4,060 adjudicated hospitalizations with available medical records occurred in 1,913 patients. Baseline characteristics of patients with at least one hospitalization event are shown in **Table 1**. Mean age was 66 years; 27% were female, 76% were White, and 18% were Black. Enrollment occurred at United States (US) Veterans Administration (VA) sites in 27%, non-VA US sites in 55%, and Canadian sites in 17%. 78% qualified for trial entry due to heart failure hospitalization, and 22% by myocardial infarction; 49% had a current or prior left ventricular ejection fraction of <40%.

**Table 1:** Baseline Characteristics of Patients With At Least One Adjudicated Hospitalization.

|  |  |
| --- | --- |
|  | n=1913 |
| Age, years | 66.3 ±13.1 |
| Female | 523 (27.3%) |
| Randomization Year |  |
| 2016-2017 | 274 (14.3%) |
| 2017-2018 | 995 (52%) |
| 2018-2019 | 644 (33.7%) |
| Region & Site |  |
| Canada | 332 (17.4%) |
| United States | 1581 (82.7%) |
| Veterans Administration | 1059 (55.4%) |
| US non-VA | 522 (27.3%) |
| Race |  |
| White | 1458 (76.2%) |
| Black | 350 (18.3%) |
| Asian | 30 (1.6%) |
| First Nation / American Indian | 16 (0.8%) |
| Other | 59 (3.1%) |
| Ethnicity |  |
| Hispanic/Latino | 136 (7.1%) |
| Non-Hispanic/Latino | 1762 (92.1%) |
| Other | 15 (0.8%) |
| Ejection fraction, % | 40.4 ±16.7 |
| NYHA Functional Class |  |
| I | 189 (13.2%) |
| II | 676 (47.0%) |
| III | 512 (35.6%) |
| IV | 60 (4.2%) |
| Body Mass Index, kg/m<sup>2</sup> | 31.7 ±8.2 |

| Qualifying Event |  |
| --- | --- |
| Heart Failure | 1496 (78.2%) |
| Myocardial Infarction | 417 (21.8%) |
| Current or Past Ejection Fraction <40% | 933 (48.8%) |
| Diabetes | 844 (44.1%) |
| Prior Heart Failure Hospitalization | 463 (24.2%) |
| History of Myocardial Infarction | 297 (15.5%) |
| History of Atrial Fibrillation | 811 (42.4%) |
| History of COPD | 444 (23.2%) |

COPD, chronic obstructive pulmonary disease; NYHA, New York Heart Association; VA, Veterans
Administration

### External validation of the Existing NLP Heart Failure Model

Out of 4,060 total hospitalizations, the CEC adjudicated 1,074 (26%) as heart failure and 2,986 as other diagnoses (**Table 2)**. The C3PO NLP model adjudicated 1486 hospitalizations (37%) as heart failure, reflecting that the externally calibrated C3PO NLP model was more permissive in adjudicating heart failure than the CEC. The C3PO NLP model adjudications demonstrated substantial agreement with the final CEC adjudication (raw agreement 87% and kappa 0.69 [95% CI 0.66-0.72]. C3PO NLP sensitivity was 94% (95% CI 92%-95%) and specificity was 84% (95% CI 83%-85%). Positive predictive value was 68% (95% CI 65%-70%) and negative predictive value was 97% (95% CI 97%-98%). Agreement between C3PO NLP and final CEC adjudication appeared to be greater in 2,195 hospitalizations in patients without history of ejection fraction <40% (kappa 0.74 vs 0.65 for 2165 hospitalizations in patients with history of ejection fraction <40%, p=0.004) and in 2,596 hospitalizations among patients age 65 or older (kappa 0.72 vs 0.66 for 1,764 hospitalizations in patients age less than 65, p=0.048), and was consistent across gender, country and type of site, race, and enrollment in the trial based on heart failure hospitalization vs myocardial infarction (**Supplemental Figure 1**). False positive C3PO NLP adjudication for heart failure was more likely in hospitalizations where the CEC adjudicated a non-heart failure cardiovascular cause (22%), particularly cardiac arrhythmia (33%), compared to pulmonary (11%) or non-cardiopulmonary causes (11%) (**Supplemental Table 1**).

**Table 2:** Heart Failure Adjudication by the C3PO NLP Model Compared to Clinical Events Committee.

|  | C3PO NLP Model |  |  |
| --- | --- | --- | --- |
| Human CEC | Heart Failure | No Heart Failure | Total |
| Heart Failure | 1009 (25%) | 65 (2%) | 1074 (26%) |
| No Heart Failure | 477 (12%) | 2509 (62%) | 2986 (74%) |
| Total | 1486 (37%) | 2574 (63%) | 4060 (100%) |
| Kappa: 0.69<br>Accuracy: 87%<br>Sensitivity: 94%<br>Specificity: 84%<br>Positive Predictive Value: 68%<br>Negative Predictive Value: 97% |  |  |  |

CEC, clinical events committee; C3PO, Community Care Cohort Project; NLP, natural language processing

A sensitivity analysis was performed in which 300 additional hospitalizations adjudicated as non-specific cardiopulmonary cause were included and considered to be heart failure. The C3PO NLP model adjudicated 199 (66%) of these cases as heart failure. With these cases included, the performance of the model decreased only slightly: accuracy was 85% and kappa 0.68, compared to 87% and 0.69 in the original cohort, respectively.

The likelihood of CEC heart failure adjudication increased across the spectrum of continuous C3PO NLP heart failure score (**Figure 2**). Just 4% of hospitalizations with NLP heart failure scores in deciles 1-5 were adjudicated as heart failure or non-specific cardiopulmonary by the CEC, compared to 89% in the top 2 deciles (in which all hospitalizations had the highest possible NLP score).

**Figure 2:**
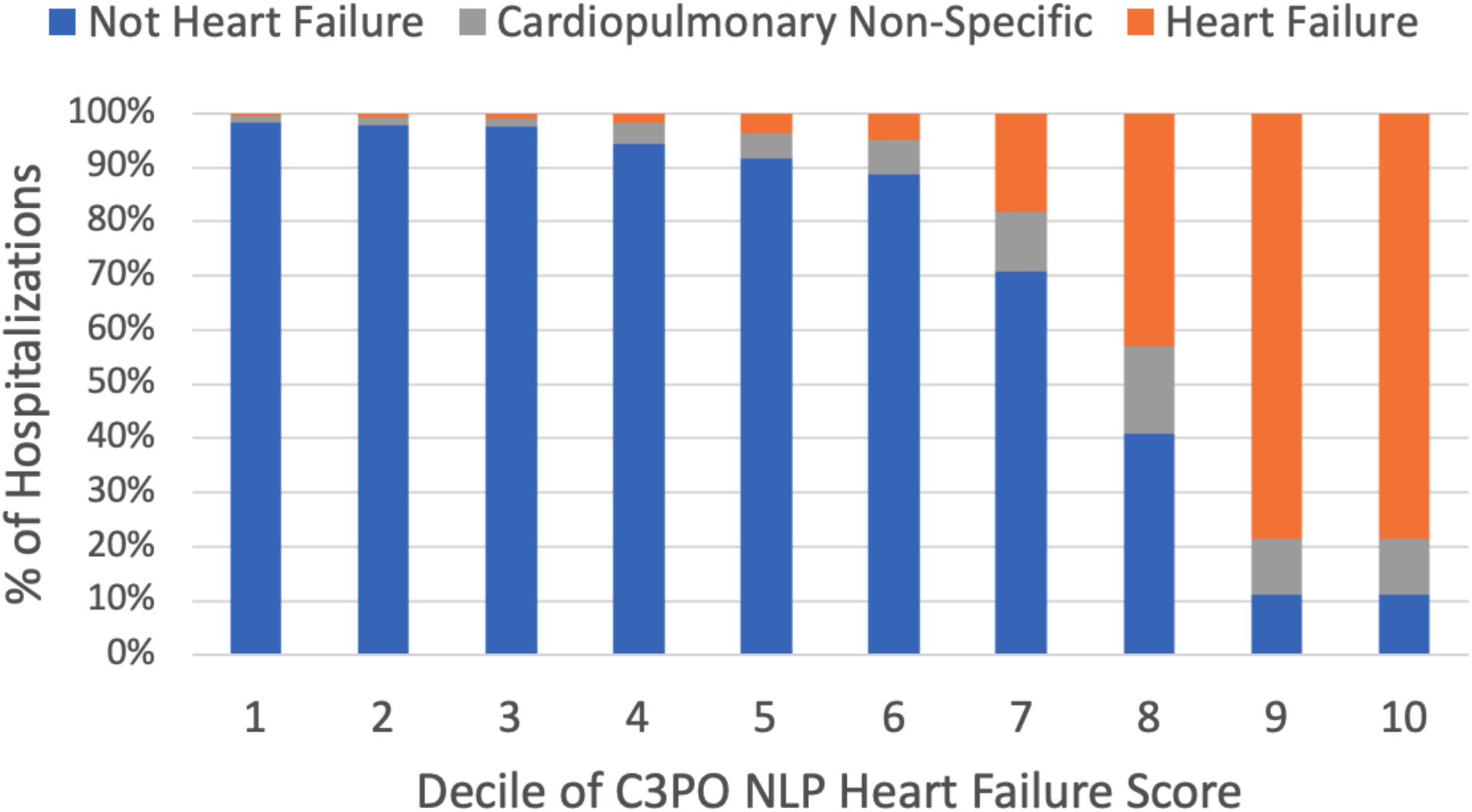
Human CEC Adjudications in Each Decile of C3PO NLP Heart Failure Score. 4360 hospitalizations were included. 960 hospitalizations with the exact highest possible C3PO NLP heart failure score were considered deciles 9 and 10. 439 hospitalizations with the identical scores just below the maximal score were considered decile 8. Deciles 1-7 include 415-430 hospitalization each.

We evaluated the accuracy of a hypothetical hybrid human + NLP strategy of using the current manual CEC process adjudication in the 20% of hospitalizations whose continuous C3PO NLP heart failure score is closest to the pre-specified threshold for heart failure (where the NLP model is most uncertain) and the NLP adjudication in the remaining 80% of hospitalizations (when the model is most confident). This strategy would have yielded 94% agreement with the final CEC adjudication (kappa 0.85 [95% CI 0.82-0.88]) while reducing the number of hospitalizations requiring manual adjudication by 80%.

### Fine-Tuning or Re-Training the NLP Model in INVESTED

Fine-tuning the C3PO NLP model or training a *de novo* NLP model using half the INVESTED hospitalizations substantially improved agreement with the gold-standard CEC adjudication. On the held-out internal validation of the remaining INVESTED hospitalizations, the C3PO+INVESTED model demonstrated kappa 0.82 (95% CI 0.77-0.86), accuracy 93%, sensitivity 81%,, specificity 98%, positive predictive value 93%, and negative predictive value 93%, relative to the gold standard of final CEC adjudication. The *de novo* INVESTED NLP model, developed on half the INVESTED hospitalization without C3PO initialization, performed similarly: kappa 0.83 (95% CI 0.79-0.87), accuracy 93%, sensitivity 84%,, specificity 97%, positive predictive value 90%, and negative predictive value 95%. Both models were superior to the original C3PO NLP model, which demonstrated a kappa statistic of 0.67 (95% CI 0.63-0.72) on the same internal validation set. The continuous NLP scores from these models (without thresholding) provided greater discrimination for the CEC-adjudicated outcome (area under the ROC curve 0.964 for C3PO+INVESTED and 0.971 for *de novo* INVESTED) than original C3PO model (0.940), indicating that training in INVESTED improved the models beyond mere recalibration.

We then investigated whether the C3PO+INVESTED and *de novo* INVESTED NLP models maintained their performance at smaller INVESTED development set sample sizes. (**Figure 3**) Fine-tuning the original C3PO NLP model with just 200-600 INVESTED examples substantially improved its agreement with the gold-standard CEC adjudication. The *de novo* INVESTED NLP model performed poorly with 200 or 400 training examples, but improved with 600 and matched the accuracy of the fine-tuned C3PO+INVESTED model at 800 examples. Both models appeared to improve modestly with additional sample size from 800 to 1954 examples.

**Figure 3:**
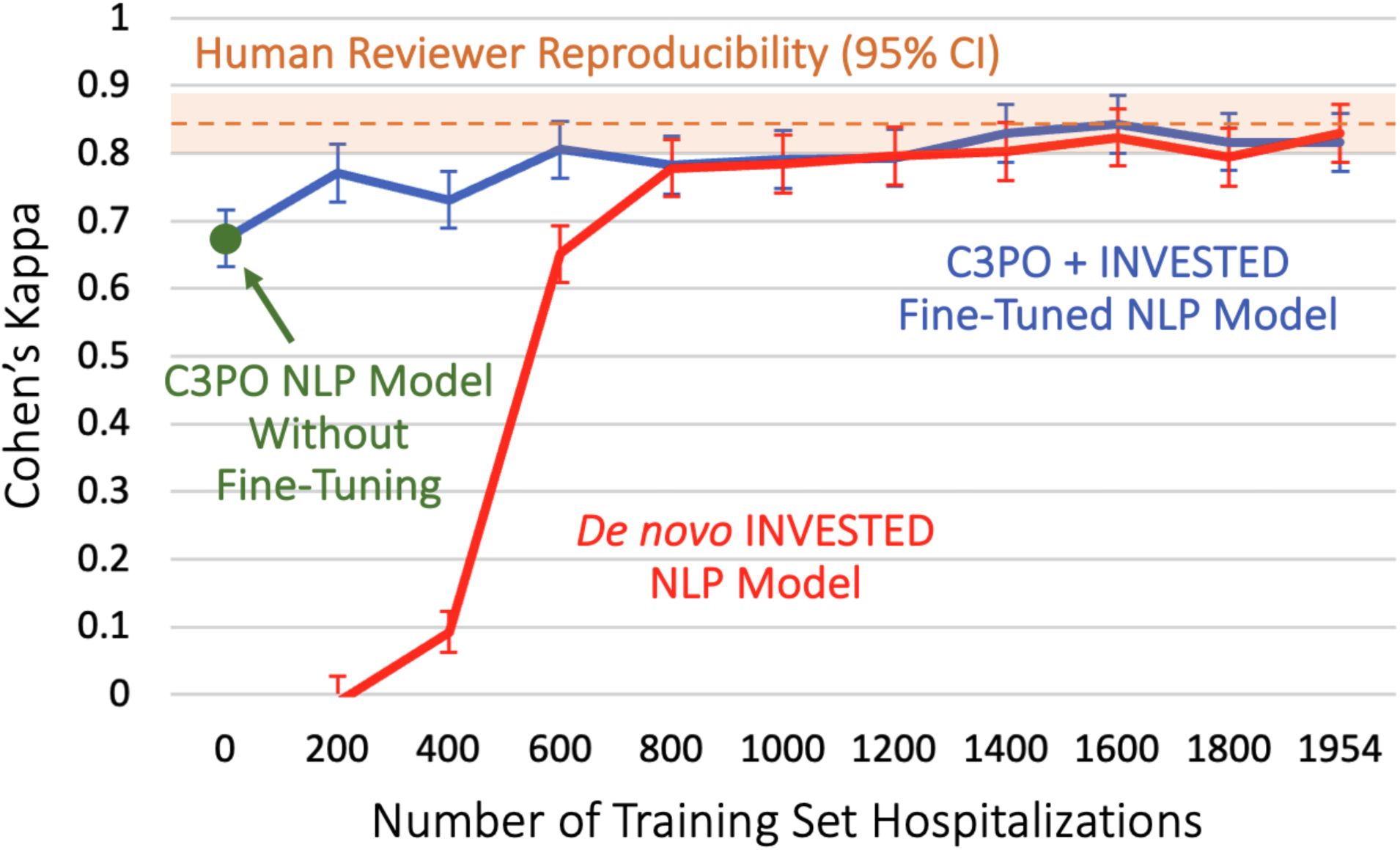
Performance of Fine-Tuned C3PO+INVESTED and *de novo* INVESTED NLP Models at Varying Training Set Sample Size. Kappa statistic for C3PO+INVESTED and *de novo* INVESTED NLP models with gold-standard CEC adjudication on a held-out internal validation set of 2106 INVESTED hospitalizations. C3PO NLP model without fine-tuning indicates external validation accuracy. Human reviewer reproducibility indicates the agreement between two independent CEC reviewers on the internal validation set. CI, confidence interval; C3PO, Community Care Cohort Project; NLP, natural language processing.

### Benchmarking NLP Adjudication to Inter-Rater Reproducibility of Two Human Reviewers

We compared the performance of the NLP models to a benchmark of human performance, defined as the inter-rater reproducibility between the two independent human reviewers that reviewed each hospitalization as part of the original INVESTED CEC. On the internal validation set, the two reviewers agreed on whether the cause of hospitalization was heart failure with accuracy 94% and kappa statistic 0.85 (95% CI 0.80-0.89). (**Figure 3**) This inter-rater reproducibility was similar to the agreement between the C3PO+ INVESTED (accuracy 93%; kappa 0.82 [95% CI 0.77-0.86]) and *de novo* INVESTED NLP models (accuracy 93%; kappa 0.83 [95% CI 0.79-0.87]), with modest differences falling within 95% confidence intervals.

## Discussion

NLP is a promising strategy for accurate and resource-efficient adjudication of clinical events directly from medical records. We previously developed an NLP model within a single-center EHR-based cohort (C3PO) that could accurately heart failure hospitalizations.^6^ We now demonstrate that this NLP model generalizes well to a multi-center clinical trial without retraining or recalibration. Specifically, C3PO NLP heart failure adjudications agreed with the gold-standard human CEC 87% of the time. Moreover, fine-tuning the C3PO NLP model or training a new NLP model in half of INVESTED hospitalizations further improved accuracy up to 93%, which approximated the inter-rater reproducibility between two human reviewers (94%). These results highlight the potential of NLP to identify clinical outcomes accurately and at scale in clinical trials and observational cohorts.

Rigorous external validation of machine learning models without retraining or recalibration is a prerequisite for meaningful implementation.^7–9^ Single center models are prone to learning artifacts such as the floor number of the heart failure ward or the name of a heart failure physician, rather than symptoms, signs, and treatment of the disease. Since these artifacts are no longer present in external datasets, models depending on them generalize poorly, a major issue in machine learning that has also been called conceptual reproducibility.^7, 8^ Our results showing substantial agreement between the C3PO NLP model and human review in a multi-center clinical trial provide reassurance that the model identifies universal evidence of heart failure rather than artifacts, and therefore can be applied to records from any North American hospital. The C3PO NLP model also proved robust to differences in document type: INVESTED dossiers were PDFs including a variety of note types (admission, progress, discharge, tables of laboratory values) whereas C3PO training data were only text format discharge summaries. Finally, generalization to INVESTED confirms that the model’s adjudications agree substantially with a gold-standard and completely independent CEC assessment central to clinical trials. Other published NLP models for heart failure have not been externally validated to our knowledge.^16, 17^

The second key finding of this study is that NLP models which were fine-tuned or *de novo* trained on a subset of INVESTED hospitalizations attained accuracy on par with human-level adjudication accuracy on the rest of INVESTED. These models agreed with the human CEC 93% of time, which was similar to the 94% inter-rater reproducibility between two expert human reviewers. These internal validation results provide optimism that future NLP models trained specifically for clinical trial event adjudication may attain human-level performance on external clinical trial datasets.

Accurate and relatively inexpensive endpoint adjudication by NLP has the potential to transform clinical trials, observational research, and quality improvement. For clinical trials, reducing CEC cost could enable larger sample sizes or trials for indications that currently lack funding, while maintaining accurate cause-specific outcomes like heart failure hospitalization which may be most strongly affected by novel therapies and are standard for regulatory approval.^5, 18^ Beyond efficacy outcomes, NLP could enable more consistent and timely reporting of adverse events in clinical trials. Further, NLP adjudication to accurately classify heart failure outcomes is broadly applicable to genomic or clinical epidemiology studies within large biobanks and registries, which have been limited by imprecise heart failure definitions using ICD codes.^6, 19^ Health policy initiatives to reduce heart failure readmissions might be more effective if targeted to hospitalizations identified by NLP rather than ICD codes.^20^ While we acknowledge that NLP models will need to be evaluated further to ensure that results are not biased in new settings, our results showing substantial agreement between NLP and CEC adjudication across multiple centers are a first step towards broad use of NLP models for these use cases.

As researchers begin to implement NLP adjudication of clinical events in future studies, combining NLP and human adjudication may be a practical approach to maintain accuracy while reducing the burden of manual adjudication. One easily implemented strategy is to manually adjudicate a subset of cases (for example, 20%) about which the model is uncertain, and to trust the NLP when it is most confident. This approach yielded 94% accuracy in our study while reducing manual adjudications by 80%.

Forthcoming innovations may improve NLP clinical event adjudication further. First, training on larger sample size datasets from multiple medical centers is likely to improve accuracy and generalizability. Performance in other machine learning models has improved with training set size in a logarithmic relationship, ^21^ which we also observed when training and fine-tuning NLP models in INVESTED. Third, generative large language models pre-trained on greater amounts of general (GPT4, LLaMa) or clinical (Med-PaLM 2) text, and with longer attention windows, ^22–24^ could be fine-tuned for event adjudication, but must be implemented carefully to maintain the security of patient health information.

Our conclusions should be interpreted in the context of limitations in the study design. First, some CEC functions, such as confirming event dates and excluding events which were not unplanned inpatient hospitalizations, were outside the scope of the C3PO NLP model’s training and therefore not evaluated. Second, in order to rigorously evaluate the C3PO NLP model on only medical record data, the NLP model was not exposed to a cover sheet containing the site investigator adjudication and brief case narrative which was seen by the CEC. Withholding this information may have led to underestimation of agreement between NLP and the CEC. Third, the method of adjudicating heart failure in the context of other cardiovascular conditions differed between INVESTED and the C3PO training set. The C3PO NLP model was trained to identify heart failure even in the presence of another primary cause of hospitalization such as arrhythmia, whereas the INVESTED CEC were asked to select a single primary diagnosis (e.g. either arrhythmia or heart failure). Agreement between the C3PO NLP model and the CEC might have been greater if the CEC had used this more permissive approach, which is more consistent with current trials in which heart failure hospitalization is a component of the primary endpoint and reflects the reality of multimorbidity.

## Conclusion

NLP is a promising strategy for accurate adjudication of clinical events at scale. In this study, the single-center C3PO NLP model for adjudication of heart failure hospitalizations generalized well to a multi-center clinical trial. C3PO NLP adjudications agreed with the gold-standard human CEC 87% of the time. Fine-tuning the C3PO NLP model or training a new NLP model further improved accuracy up to 93%, which was similar to human reviewer reproducibility. These results highlight the potential of NLP to identify clinical outcomes accurately and scalably in clinical trials and observational cohorts.

### Data Availability Statement

Deidentified participant data from the INVESTED trial will be available on the NHLBI Biolincc website to qualified researchers as per NHBLI guidelines.

### Funding

The INVESTED trial was funded by the National Heart, Lung, and Blood Institute. Dr. Cunningham is supported by the KL2/Harvard Catalyst Medical Research Investigator Training program and the American Heart Association (23CDA1052151). Dr. Lau is supported by the NIH (K23-HL159243) and the American Heart Association (853922). Dr. Ellinor is supported by the NIH (1R01HL092577, K24HL105780), AHA (18SFRN34110082), Foundation Leducq (14CVD01), and by MAESTRIA (965286). Dr. Ho is supported by the NIH (R01 HL134893, R01 HL140224, R01HL160003, and K24 HL153669).

## References

1. Ziaeian B, Fonarow GC. The Prevention of Hospital Readmissions in Heart Failure. Emerg Trends Curr Controv Heart Fail. 2016;58:379–385.

2. Hicks Karen A., Mahaffey Kenneth W., Mehran Roxana, Nissen Steven E., Wiviott Stephen D., Dunn Billy, Solomon Scott D., Marler John R., Teerlink John R., Farb Andrew, Morrow David A., Targum Shari L., Sila Cathy A., Hai Mary T. Thanh, Jaff Michael R., Joffe Hylton V., Cutlip Donald E., Desai Akshay S., Lewis Eldrin F., Gibson C. Michael, Landray Martin J., Lincoff A. Michael, White Christopher J., Brooks Steven S., Rosenfield Kenneth, Domanski Michael J., Lansky Alexandra J., McMurray John J.V., Tcheng James E., Steinhubl Steven R., Burton Paul, Mauri Laura, O’Connor Christopher M., Pfeffer Marc A., Hung H.M. James, Stockbridge Norman L., Chaitman Bernard R., Temple Robert J. 2017 Cardiovascular and Stroke Endpoint Definitions for Clinical Trials. Circulation. 2018;137:961–972.

3. Strom Jordan B., Faridi Kamil F., Butala Neel M., Zhao Yuansong, Tamez Hector, Valsdottir Linda R., Brennan J. Matthew, Shen Changyu, Popma Jeffrey J., Kazi Dhruv S., Yeh Robert W. Use of Administrative Claims to Assess Outcomes and Treatment Effect in Randomized Clinical Trials for Transcatheter Aortic Valve Replacement. Circulation. 2020;142:203–213.

4. Danaei G. Causal Analyses of Nested Case-Control Studies for Comparative Effectiveness Research. PCORI Public Prof Res Rep. 2021;

5. Cowie MR, Blomster JI, Curtis LH, Duclaux S, Ford I, Fritz F, Goldman S, Janmohamed S, Kreuzer J, Leenay M, Michel A, Ong S, Pell JP, Southworth MR, Stough WG, Thoenes M, Zannad F, Zalewski A. Electronic health records to facilitate clinical research. Clin Res Cardiol. 2017;106:1–9.

6. Cunningham Jonathan W., Singh Pulkit, Reeder Christopher, Lau Emily S., Khurshid Shaan, Wang Xin, Ellinor Patrick T., Lubitz Steven A., Batra Puneet, Ho Jennifer E. Natural Language Processing for Adjudication of Heart Failure in the Electronic Health Record. JACC Heart Fail [Internet]. [cited 2023 May 9];0. Available from: https://doi.org/10.1016/j.jchf.2023.02.012

7. McDermott MBA, Wang S, Marinsek N, Ranganath R, Foschini L, Ghassemi M. Reproducibility in machine learning for health research: Still a ways to go. Sci Transl Med. 2021;13:eabb1655.

8. Barak-Corren Y, Chaudhari P, Perniciaro J, Waltzman M, Fine AM, Reis BY. Prediction across healthcare settings: a case study in predicting emergency department disposition. Npj Digit Med. 2021;4:169.

9. Yang J, Soltan AAS, Clifton DA. Machine learning generalizability across healthcare settings: insights from multi-site COVID-19 screening. Npj Digit Med. 2022;5:69.

10. Vardeny O, Udell JA, Joseph J, Farkouh ME, Hernandez AF, McGeer AJ, Talbot HK, Bhatt DL, Cannon CP, Goodman SG, Anand I, DeMets DL, Temte J, Wittes J, Nichol K, Yancy CW, Gaziano JM, Cooper LS, Kim K, Solomon SD. High-dose influenza vaccine to reduce clinical outcomes in high-risk cardiovascular patients: Rationale and design of the INVESTED trial. Am Heart J. 2018;202:97–103.

11. Vardeny O, Kim K, Udell JA, Joseph J, Desai AS, Farkouh ME, Hegde SM, Hernandez AF, McGeer A, Talbot HK, Anand I, Bhatt DL, Cannon CP, DeMets D, Gaziano JM, Goodman SG, Nichol K, Tattersall MC, Temte JL, Wittes J, Yancy C, Claggett B, Chen Y, Mao L, Havighurst TC, Cooper LS, Solomon SD, INVESTED Committees and Investigators. Effect of High-Dose Trivalent vs Standard-Dose Quadrivalent Influenza Vaccine on Mortality or Cardiopulmonary Hospitalization in Patients With High-risk Cardiovascular Disease: A Randomized Clinical Trial. JAMA. 2021;325:39–49.

12. Artifex Software. Ghostscript. [Internet]. [cited 2023 May 9];Available from: www.ghostscript.com

13. Kay A. Tesseract: an open-source optical character recognition engine. Linux J. 2007;2007:2.

14. Khurshid S, Reeder C, Harrington LX, Singh P, Sarma G, Friedman SF, Di Achille P, Diamant N, Cunningham JW, Turner AC, Lau ES, Haimovich JS, Al-Alusi MA, Wang X, Klarqvist MDR, Ashburner JM, Diedrich C, Ghadessi M, Mielke J, Eilken HM, McElhinney A, Derix A, Atlas SJ, Ellinor PT, Philippakis AA, Anderson CD, Ho JE, Batra P, Lubitz SA. Cohort design and natural language processing to reduce bias in electronic health records research. Npj Digit Med. 2022;5:47.

15. Li Y, Wehbe RM, Ahmad FS, Wang H, Luo Y. Clinical-Longformer and Clinical-BigBird: Transformers for long clinical sequences [Internet]. 2022;Available from: https://arxiv.org/abs/2201.11838

16. Goto S, Homilius M, John JE, Truslow JG, Werdich AA, Blood AJ, Park BH, MacRae CA, Deo RC. Artificial intelligence-enabled event adjudication: estimating delayed cardiovascular effects of respiratory viruses. medRxiv. 2020;2020.11.12.20230706.

17. Ambrosy AP, Parikh RV, Sung SH, Narayanan A, Masson R, Lam P-Q, Kheder K, Iwahashi A, Hardwick AB, Fitzpatrick JK, Avula HR, Selby VN, Shen X, Sanghera N, Cristino J, Go AS. A Natural Language Processing–Based Approach for Identifying Hospitalizations for Worsening Heart Failure Within an Integrated Health Care Delivery System. JAMA Netw Open. 2021;4:e2135152–e2135152.

18. Mentz RJ, Anstrom KJ, Eisenstein EL, Sapp S, Greene SJ, Morgan S, Testani JM, Harrington AH, Sachdev V, Ketema F, Kim D-Y, Desvigne-Nickens P, Pitt B, Velazquez EJ, TRANSFORM-HF Investigators. Effect of Torsemide vs Furosemide After Discharge on All-Cause Mortality in Patients Hospitalized With Heart Failure: The TRANSFORM-HF Randomized Clinical Trial. JAMA. 2023;329:214–223.

19. Aragam KG, Chaffin M, Levinson RT, McDermott G, Choi SH, Shoemaker MB, Haas ME, Weng L-C, Lindsay ME, Smith JG, Newton-Cheh C, Roden DM, London B, null null, Wells QS, Ellinor PT, Kathiresan S, Lubitz SA, Bloom HL, Dudley SC, Shalaby AA, Weiss R, Gutmann R, Saba S. Phenotypic Refinement of Heart Failure in a National Biobank Facilitates Genetic Discovery. Circulation. 2019;139:489–501.

20. Wadhera RK, Joynt Maddox KE, Wasfy JH, Haneuse S, Shen C, Yeh RW. Association of the Hospital Readmissions Reduction Program With Mortality Among Medicare Beneficiaries Hospitalized for Heart Failure, Acute Myocardial Infarction, and Pneumonia. JAMA. 2018;320:2542–2552.

21. Mahajan D, Girshick R, Ramanathan V, He K, Paluri M, Li Y, Bharambe A, Maaten L van der. Exploring the Limits of Weakly Supervised Pretraining. 2018;

22. Nori H, King N, McKinney SM, Carignan D, Horvitz E. Capabilities of gpt-4 on medical challenge problems. ArXiv Prepr ArXiv230313375. 2023;

23. Li Y, Li Z, Zhang K, Dan R, Zhang Y. ChatDoctor: A Medical Chat Model Fine-tuned on LLaMA Model using Medical Domain Knowledge. 2023;

24. Singhal K, Azizi S, Tu T, Mahdavi SS, Wei J, Chung HW, Scales N, Tanwani A, Cole-Lewis H, Pfohl S, Payne P, Seneviratne M, Gamble P, Kelly C, Scharli N, Chowdhery A, Mansfield P, Arcas BA y, Webster D, Corrado GS, Matias Y, Chou K, Gottweis J, Tomasev N, Liu Y, Rajkomar A, Barral J, Semturs C, Karthikesalingam A, Natarajan V. Large Language Models Encode Clinical Knowledge. 2022;

