## Supplemental Figure 1 for "Natural Language Processing for Adjudication of Heart Failure Hospitalizations in a Multi-Center Clinical Trial"

**Table of Contents**

Supplemental Table 1: Rate of NLP Heart Failure by True CEC Adjudication……………………………2

Supplemental Figure 1: Agreement Between NLP and Human CEC Heart Failure Adjudications in Key Subgroups of Patients ………………………………………………………………………………3

**Supplemental Table 1: Rate of NLP Heart Failure by True CEC Adjudication**

| **CEC Adjudication** | Total Hospitalizations | NLP HF Hospitalizations | % NLP HF |
|---|---|---|---|
| Heart Failure | 1,074 | 1,009 | 94% |
| Cardiopulmonary Non-Specific | 300 | 199 | 66% |
| Non-HF Cardiovascular Causes | 1375 | 297 | 22% |
| Pulmonary | 290 | 31 | 11% |
| Non-Cardiopulmonary | 1,305 | 147 | 11% |
| Unknown | 16 | 2 | 13% |

HF, heart failure; NLP, natural language processing.

**Supplemental Figure 1: Agreement Between NLP and Human CEC Heart Failure Adjudications in Key Subgroups of Patients**

| Subgroup | | Hospitalizations | Kappa (95% CI) | $P_{Interaction}$ |
|---|---|---|---|---|
| Age | <65 | 1,764 | 0.66 (0.61-0.71) | 0.05 |
| | 65+ | 2,596 | 0.72 (0.68-0.76) | |
| Gender | Female | 1,121 | 0.71 (0.65-0.77) | 0.48 |
| | Male | 3,147 | 0.69 (0.65-0.72) | |
| Race | White | 2,502 | 0.70 (0.66-0.73) | 0.48 |
| | Black | 107 | 0.67 (0.61-0.74) | |
| Site | US Non-VA | 2,502 | 0.68 (0.64-0.72) | |
| | US VA | 1,289 | 0.74 (0.68-0.79) | 0.08 |
| | Canada | 569 | 0.66 (0.58-0.74) | 0.74 |
| Qualifying by HF | Yes | 3,598 | 0.68 (0.65-0.72) | 0.67 |
| | No | 762 | 0.67 (0.6-0.74) | |
| Current or Prior EF <40% | Yes | 2,165 | 0.65 (0.61-0.69) | 0.003 |
| | No | 2,195 | 0.74 (0.70-0.78) | |

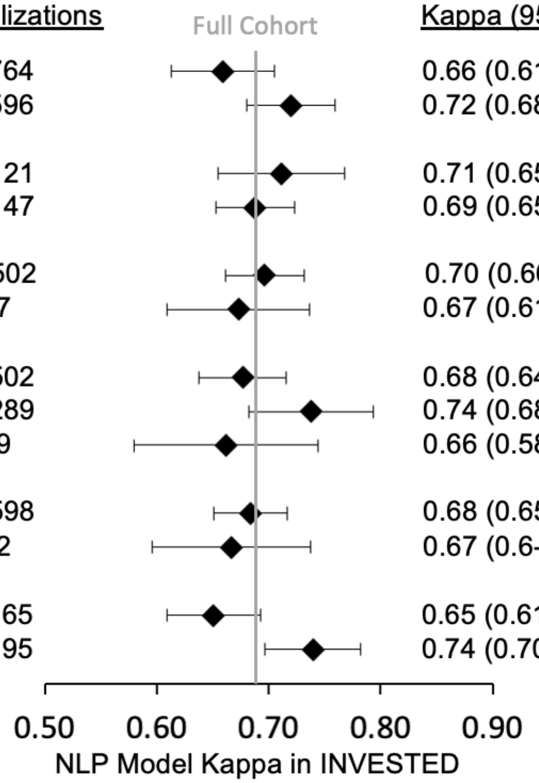


Interaction p-value evaluates the null hypothesis that the kappa statistic does not differ between subgroups. CI, confidence interval. EF, ejection fraction; HFH, heart failure hospitalization; VA, Veterans Administration.
